# Geospatial and Socioeconomic Analysis of Direct Primary Care Practices in the United States

**DOI:** 10.1101/2025.09.20.25336249

**Authors:** Juan P. Carabeo-Nieva, Oscar A. Lugo Capera

**Affiliations:** Universidad Central del Caribe (Bayamon, PR)

## Abstract

**Background:** Current literature on direct primary care (DPC) is largely composed of opinion-based arguments that the model may exacerbate healthcare inequity. Objective data are needed to assess how DPC practices are distributed and whether they contribute to disparities in access to care.

**Methods:**

1. Visited DPC Frontier website (https://mapper.dpcfrontier.com).
2. Extracted clinic information from the website using a custom web-socket script (ws.py) that requested each entry using its practice-key (scraped from raw html).
3. Duplicate entries removed by R script (No-Dups.R) unique.data.frame() (unique rows) (Data_Columns = “Postal”,“City”,“Region”,“Name”,“Lat”,“Lng”,“Website”)
3. Clinic data was then merged (by zipcode) with the metadata from the zipcodeR database and the Rural-Urban Commuting Area Codes (RUCA) coding table from the US Department of Agriculture to produce the Mergd-with-RUCA.csv spreadsheet (a comprehensive geospatial database of DPC practice locations and their associated demographic and socioeconomic characteristics).
4. Data Analysis: Data-Analysis-Recommendations.pdf Sections 2,4, and 5 (authored by Sr. Lugo Capera)

**Results:** DPC practices were more common in urban and suburban zip codes, positively correlated with higher total housing units, lower occupancy rates, and lower median household incomes. Family medicine was more prevalent in lower-income zip codes, while specialties such as dermatology and cardiology clustered in middle- and higher-income areas.

**Discussion:** While DPC practices appear to favor more commercial or suburban settings, their presence in lower-income zip codes suggests potential to serve populations with limited access to traditional care. However, specialty care appears less equitably distributed.

**Conclusion:** The GINI coefficient of 0.22 for DPC practice distribution indicates modest inequality, with most zip codes hosting one or no DPC practices. While geographic access to primary DPC appears relatively even, disparities in specialty DPC and potential quality differences merit further investigation.

## Introduction

Direct primary care (DPC) is a healthcare delivery model in which patients pay a recurring membership fee directly to a physician or clinic, bypassing traditional insurance-based reimbursement. Proponents argue that this model improves access, enhances the patient-physician relationship, and reduces administrative burden.1 However, critics have raised concerns that DPC may worsen health inequities by prioritizing wealthier or urban populations and reducing access for vulnerable groups.^2,3^

Despite growing interest, current literature on DPC is primarily qualitative or opinion-based, with limited empirical data analyzing the distribution and accessibility of DPC practices. To date, few studies have investigated the geospatial and socioeconomic characteristics of DPC practice locations, or the implications for health equity.4

This study aims to address this gap by using publicly available datasets to quantify the geographic and demographic distribution of DPC practices across the United States, offering an early empirical perspective on whether DPC contributes to or mitigates healthcare disparities.

## Methods

Data Sources and Integration

Three primary datasets were used:

DPC Frontier Mapper – a crowd-sourced and regularly updated map of DPC clinics across the United States.^5^

zipcodeR R package – an open-source resource containing over 40 demographic and housing-related variables for U.S. zip codes.^6^

Rural-Urban Commuting Area (RUCA) codes – developed by the USDA to classify U.S. census tracts by urbanicity and commuting patterns.^7^

These datasets were merged based on zip code to associate DPC practice locations with socioeconomic and geographic characteristics.

Variables and Statistical Analysis

Key variables analyzed included:

Urban-rural classification (RUCA code)

Median household income

Total and occupied housing units

Specialty of practice (family medicine, cardiology, dermatology, etc.)

Descriptive statistics were used to characterize distribution patterns. Correlations were assessed using Pearson coefficients. Inequality in distribution was measured using the GINI coefficient, calculated based on DPC practices per zip code.

All analyses were conducted in R (v4.3.0).

## Results

### Geographic and Urban-Rural Distribution

DPC practices were predominantly located in urban and suburban areas. Very few were present in rural zip codes with RUCA codes >6, indicating a skew toward more densely populated regions.

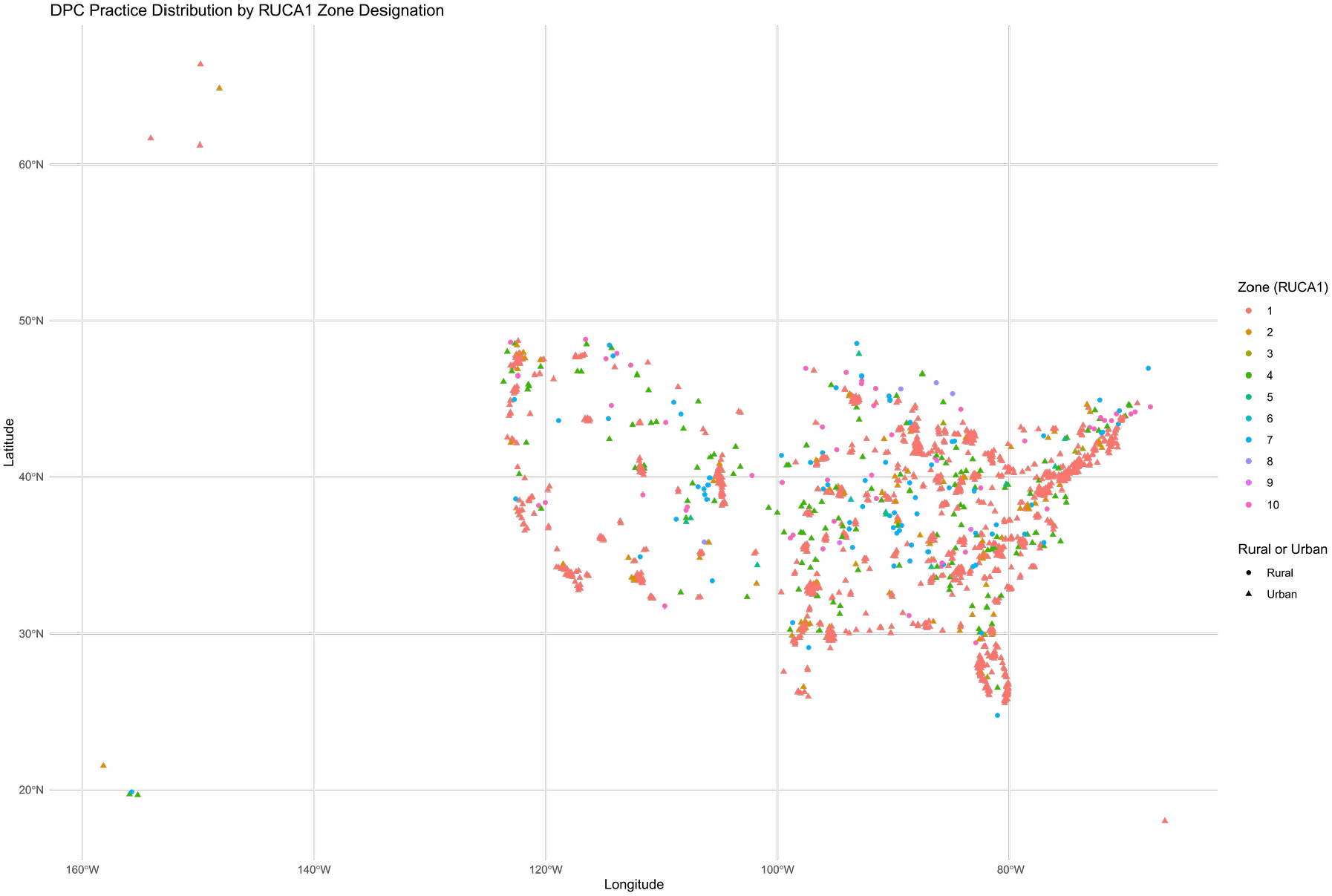

Socioeconomic Correlates

There was a positive correlation between the presence of a DPC practice and:

Total housing units (r = 0.47)

Lower housing occupancy rates (r = –0.36)

Lower median household income (r = –0.22)

This finding suggests that while practices are located in more commercialized or populated areas, they may be serving areas with economic disadvantage.

### Specialty Distribution

Family medicine was the most commonly represented specialty across all areas, particularly in lower-income zip codes. In contrast, specialties such as dermatology and cardiology were only found in middle- and higher-income areas, and in zip codes with higher commercial density.

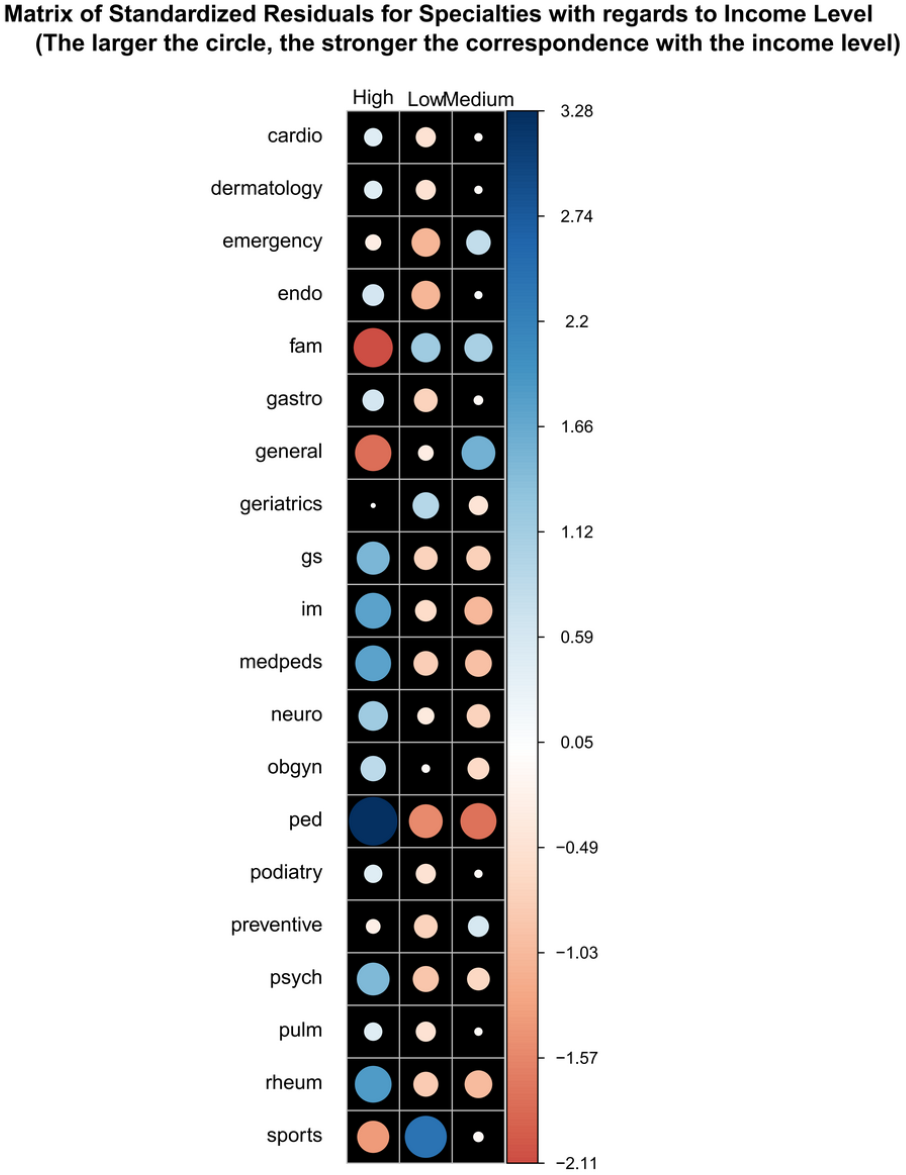

### Inequality of Distribution

The GINI coefficient for DPC practice distribution across zip codes was 0.22, indicating relatively modest inequality. Most zip codes had no DPC practices; those that did usually had one, with very few having multiple.

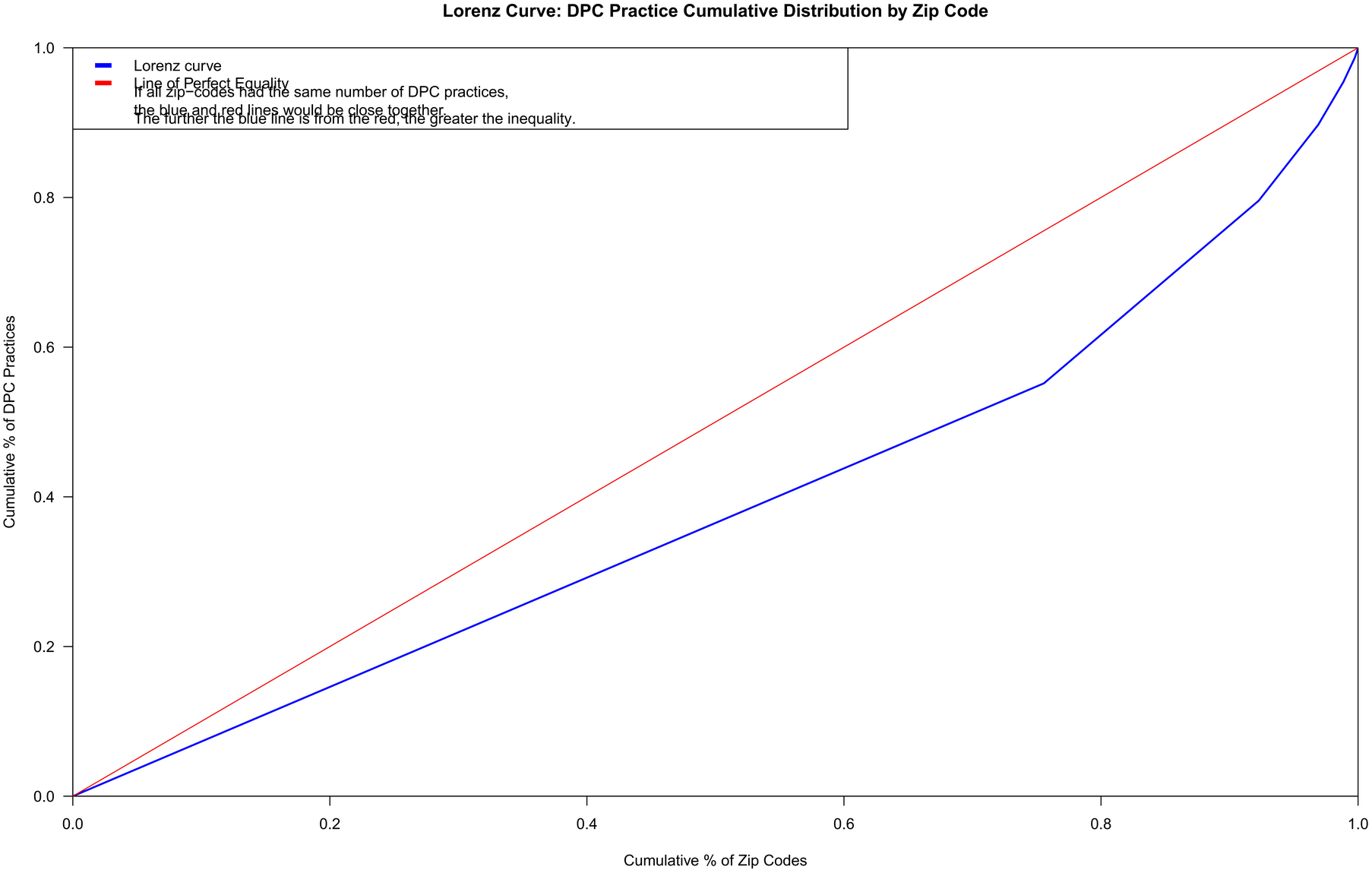

## Discussion

This analysis reveals that DPC practices are unevenly distributed, favoring urban and suburban settings. However, their presence in lower-income zip codes complicates the narrative that DPC is only for the affluent. These findings suggest DPC may fill care gaps in under-resourced urban areas where traditional primary care is less available. Nonetheless, specialty care within the DPC model appears far less equitably distributed. Cardiology and dermatology practices were absent from low-income zip codes, highlighting a persistent disparity in specialty access. The modest GINI coefficient indicates a more even geographic spread than might be expected, especially when compared to traditional healthcare infrastructure, which has long been concentrated in wealthier urban cores.^8^ However, the true measure of equity requires assessing not just distribution but also quality and affordability of care delivered. Further research is needed to evaluate outcomes such as patient satisfaction, clinical quality, and cost-effectiveness, particularly in comparison to traditional fee-for-service models.

## Conclusion

DPC practices are more common in urbanized areas but are not limited to wealthy zip codes. The relatively low GINI coefficient suggests modest inequality in geographic distribution. While primary care appears to be fairly accessible through DPC, disparities remain for specialty care.

Future studies should explore the quality and affordability of DPC services and whether the model improves outcomes for underserved populations or simply shifts care for those with existing access.

## Supporting information

Custom_Database

## Data Availability

All data produced in the present study are available upon reasonable request to the authors.

https://mapper.dpcfrontier.com

## Notes

### Competing Interest Statement

The authors have declared no competing interest.

### Funding Statement

This study did not receive any funding.

## References

1. Eskew P. Direct primary care: Practice distribution and cost across the nation. J Am Board Fam Med. 2018;31(4):588–589. doi:10.3122/jabfm.2018.04.170388

2. Phillips RL Jr, Liaw WR, Ko M, et al. Specialty and geographic distribution of the physician workforce: What influences medical student & resident choices? Acad Med. 2009;84(2):222–226. doi:10.1097/ACM.0b013e318193eb53

3. Kullgren JT. Opinion: The trouble with direct primary care. Health Affairs Blog. Published March 2018. Accessed August 21, 2025. https://www.healthaffairs.org/do/10.1377/forefront.20180301.830206

4. Neutze D, DeVoe J. Is direct primary care the solution to inequity in primary care access? Ann Fam Med. 2017;15(4):303–305. doi:10.1370/afm.2116

5. DPC Frontier Mapper. Accessed August 10, 2025. https://mapper.dpcfrontier.com/

6. zipcodeR package. GitHub repository. https://github.com/gavinrozzi/zipcodeR

7. USDA Economic Research Service. Rural-Urban Commuting Area Codes. https://www.ers.usda.gov/data-products/rural-urban-commuting-area-codes

8. Gaskin DJ, Dinwiddie GY, Chan KS, McCleary R. Residential segregation and the availability of primary care physicians. Health Serv Res. 2012;47(6):2353–2376. doi:10.1111/j.1475-6773.2012.01417.x

